# More Than Results: A Qualitative Study on the Role of Person-Centered Genetic Counseling in Parkinson’s Disease Research

**DOI:** 10.64898/2026.06.03.26354465

**Authors:** Jennifer Verbrugge, Katie Fiallos, Lola Cook, Mandy Miller, Katharine J. Head

**Affiliations:** Department of Medical and Molecular Genetics. Indiana University School of Medicine, 410 W. 10th St., Indianapolis, IN, USA; Department of Communication, 425 University Blvd, Indiana University, Indianapolis, IN, USA

**Keywords:** Parkinson’s disease, genetic counseling, results disclosure, *LRRK2*, *GBA1*

## Abstract

As genetic testing becomes increasingly integrated into Parkinson’s disease (PD) research, including targeted testing for variants in *LRRK2* and *GBA1*, the return of individual research results is becoming more common. However, limited qualitative data exists regarding how research participants experience genetic results disclosure and post-test genetic counseling in PD research settings. We conducted semi-structured qualitative interviews with participants (n=13) enrolled in the Parkinson’s Precision Medicine Initiative (formerly Parkinson’s Progression Markers Initiative; PPMI) who had received PD-related genetic test results and post-test genetic counseling. Interviews were conducted 1–3 weeks following result disclosure and analyzed using thematic analysis with a primarily deductive coding approach informed by study aims and inductive identification of emergent themes. Four primary themes were identified: (1) personal connection and motivations for participation, (2) centrality of result disclosure and information preferences, (3) emotional experiences and support needs, and (4) communication quality and alignment with participant needs. Overall, our findings underscore the importance of person-centered genetic counseling within PD research. As return of genetic and biomarker results in research and clinical trial contexts expand, thoughtful integration of relational, informational, and communication-focused practices will be essential to support participant engagement and trust.

## Background

Parkinson’s disease (PD) is a common neurodegenerative disorder with both genetic and environmental contributors. Although most PD cases are considered sporadic, pathogenic variants in genes such as *LRRK2* and *GBA1* represent the most frequently known genetic risk factors and have become increasingly relevant as genotype-specific clinical trials emerge. As a result, genetic testing for PD-related variants is expanding to include risk stratification, trial eligibility, and research participation [1].

Despite the growing incorporation of genetic testing into PD research and the increasing return of individual genetic results, limited data have been available on how individuals with PD or those at-risk experience research genetic result disclosure and post-test genetic counseling, including their informational needs, emotional responses, and communication preferences. This gap is notable given that learning one’s genetic status may have emotional and behavioral implications, particularly in a disease with variable gene penetrance and currently limited disease-modifying therapies. Prior work in other disease contexts suggests that preferences for education, support, and communication during return of results vary meaningfully across participants, underscoring the importance of characterizing participant experiences in research settings [2;3]. The Parkinson’s Precision Medicine Initiative (formerly the Parkinson’s Progression Markers Initiative; PPMI) is a large, international, longitudinal observational study designed to identify biomarkers of PD diagnosis and progression. PPMI has been central to advancing understanding of PD by recruiting individuals with PD and those at elevated risk and combining targeted genetic testing with biomarker and other assessments. Within PPMI, individuals meeting specific eligibility criteria have undergone targeted testing for pathogenic variants in *LRRK2* and *GBA1*, with results returned through structured disclosure processes that include genetic counseling by certified genetic counselors [4–6].

To begin addressing some of these questions regarding participant experiences, Verbrugge et al [6] provided quantitative data on participant-reported outcomes following genetic counseling in a large PD research genetics cohort. Specifically, they evaluated the psychological impact of receiving PD-related genetic test results and satisfaction with the genetic counseling process among individuals with PD and those at increased risk who underwent targeted *LRRK2* and *GBA1* testing through PPMI. Overall, participant-reported outcomes indicated that genetic testing and result disclosure are generally well received, with most participants reporting favorable psychological impact and high satisfaction with genetic counseling. The current qualitative study was designed to gather a more in-depth understanding of the genetic testing and counseling experiences and potential needs among PPMI research participants to guide future research efforts.

## METHODS

### Participants

Individuals enrolled in PPMI who met eligibility criteria for PD-related genetic testing, including those with PD or elevated genetic risk, underwent targeted testing for *LRRK2* and *GBA1* variants. Genetic results were then disclosed via genetic counseling by certified genetic counselors over telephone or video conference. Participants who completed the genetic counseling session were subsequently invited to participate in this semi-structured qualitative interview study.

Purposive sampling was used to enroll individuals with variation in sex, PD status, and genetic result to enhance depth and diversity of perspectives. During the qualitative study period, PPMI shifted from enrolling participants with and without PD to enrolling only those without PD, which limited the number of participants with PD. Sample size was guided by the concept of information power [7]. Interviews were conducted between March 2022 and April 2023, and participants received compensation ($30 gift card). The study was approved by the Indiana University Institutional Review Board, and all participants provided informed consent.

### Interviews

An interview guide was developed by three genetic counselors (JV, LC, and MM) and an expert in health communications research (KH). The interview guide encompassed questions pertaining to domains associated with the return of genetic research results and genetic counseling, with particular attention to informational and communication needs, emotional aspects, and participant expectations. A copy of the interview guide is available in the Supplement. We conducted interviews 1-3 weeks after the genetic counseling session, with each interview lasting 30-45 minutes. Three genetic counselors conducted the interviews (JV, LC, and MM). The genetic counselor who performed the interview was not the same genetic counselor who disclosed results and provided genetic counseling to the participant. The interviews were performed remotely and recorded through a Zoom/Microsoft Teams video meeting, transcribed, and analyzed.

### Data Analysis

Thematic analysis was used to identify themes in qualitative interviews, as described by Braun and Clark [8]. A primarily deductive coding approach was used, guided by the interview structure and research questions, while inductive coding was included to allow for unexpected themes to emerge directly from participants’ perspectives. All recorded interviews were transcribed and uploaded to Dedoose version 10.0.25 coding platform. Four coders (JV, KF, LC, and MM) independently reviewed and line-by-line coded transcripts, met to develop and refine a shared codebook, and applied the finalized codebook to remaining transcripts. Themes were identified through iterative consensus discussions. All coders reviewed the final analysis and agreed upon the final themes and exemplary quotations that are presented in the results section.

### Data Sharing

Raw data of transcribed interviews will not be publicly available because this data sharing could compromise anonymity, and participants did not provide consent for this data sharing. The consent document informed participants that their interview data could be used for publication purposes.

## RESULTS

Interviews were conducted with 13 participants (one interview included a participant’s spouse who had also taken part in the genetic counseling session) with a median age of 69 years old, of whom three had a diagnosis of PD, and ten were at increased risk for PD. Three participants received genetic counseling for positive genetic test results, and the remaining ten participants had negative results (see Table 1 for full participant characteristics). Of those with positive results, one participant had a diagnosis of PD, and the other two were unaffected at the time of genetic testing and counseling. Analysis of codes and subcodes led to identification of four themes that focused on return of research genetic results and genetic counseling: 1) personal connection and motivation for participation, 2) centrality of result disclosure and information preferences, 3) emotional experiences and support needs in result disclosure, and 4) communication quality and alignment with participant needs.

**Table 1.**
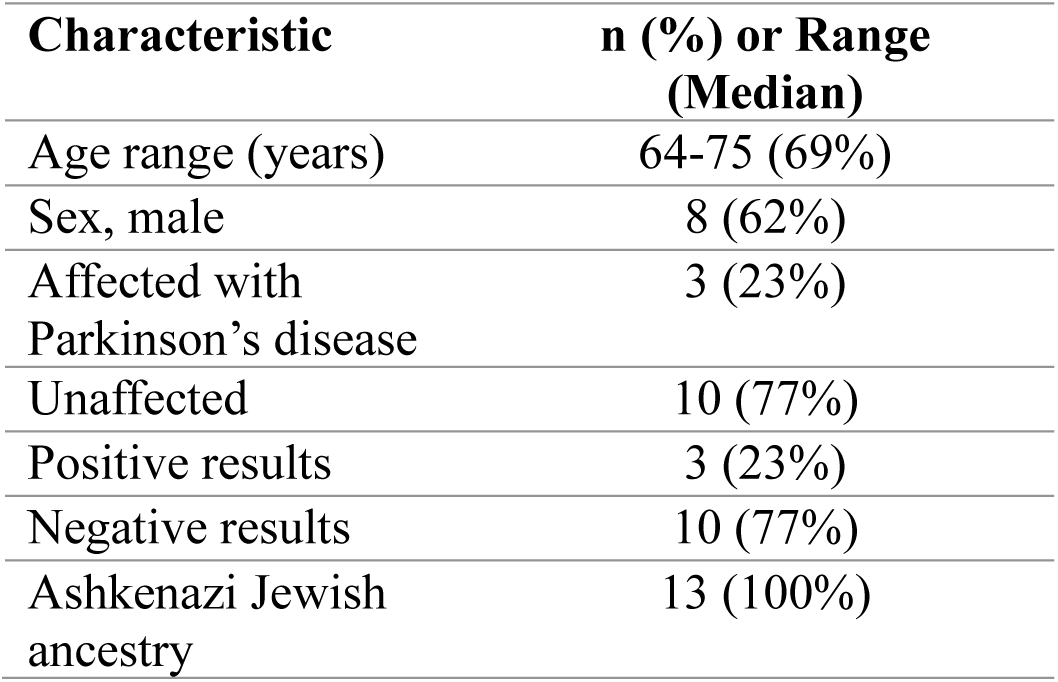
Participant Characteristics (n = 13)

### Theme 1: Personal Connection and Motivations for Participation

The interview did not include specific questions about why individuals chose to participate in the genetic testing portion of the study. Nevertheless, some participants raised this topic, and often their personal and family connection to PD were primary motivators.

> ***Participant 2 (age 60-64, unaffected male, negative results):*** *“But, you know, the reason I did this was I have two parents who -- one who died of Parkinson’s-related illness, the other had Parkinson’s. I have a partner who has REM sleep disturbance. So, you know, I have a real interest in promoting the research and that’s why I did it.”*

For some, this idea of a personal connection extended beyond just a link to PD. For example, one participant mentioned that the personal touch of providing genetic counseling was a motivator for them to participate.

> ***Participant 1 (age 65-69, affected male, negative results):*** *“Well, I was actually kind of thankful to actually have that session more than just a letter in the mail with the test result. So, the fact that there was an interpersonal component to your overall process was quite welcomed, and I think the fact that that was there, and part of the process was part of the incentive, or the inducement for me to go ahead and participate.”*

As shown here, participation in the research was personal for some, making individualized counseling especially important.

### Theme 2: Centrality of Result Disclosure and Information Preferences

Continuing the overarching idea of research being personal, participants highlighted their desire to receive information about themselves from the study. Even though those who raised the topic of why they chose to enroll in the study often talked about family and promoting research as key motivators, many participants emphasized that receiving their results was the most important part of the session, underscoring the intrinsic value participants placed on receiving individual research results.

> ***Participant 9 (age 65-69, unaffected male, negative results):*** *“I mean, the results. Truly, all I really wanted to hear was the results. Everything was fill-in, fluff, icing, whatever you want to call it. The results are all that mattered first.”*

To this point, some participants expressed an unmet desire for an even broader return of personalized information, including genetic findings beyond PD or other available study data, such as the results of the smell test also done through the study.

> ***Participant 11 (age 65-69, unaffected male, negative results):*** *“I expected to get a little more overall information about my general health…I mean, as long as you guys were doing the tests and I’m sure the information is generated, it would have been nice to get the feedback about, well, you know, there was nothing interesting there relating to Parkinson’s but you did have a marker for, increased risk of heart disease or whatever.*

This desire for information extended beyond study results to wanting more general information that they could potentially apply individually.

> ***Participant 1 (age 65-69, unaffected male, negative results):*** *“I guess I would have liked to have heard a little bit more about environmental factors, but then again, my sense was that it wasn’t what you guys were focused on, so I didn’t pursue that line of questions.”*
>
> ***Participant 4 (age 70-74, unaffected female, GBA1 positive result):*** *“Yeah. It would’ve been nice to find out how to recognize symptoms if I am getting [Parkinson’s disease].”*

Aside from what information participants valued, they also answered questions about ways they prefer to receive information prior to post-test genetic counseling. In this study, pre-test genetic counseling was available but not required, and interview participants generally agreed with keeping this optional. However, many participants reported they were unaware of the option for pre-test counseling. A few shared they would have requested pre-test genetic counseling had they known it was available, but most felt they did not need it.

> ***Participant 7 (age 65-69, unaffected male, negative results):*** *“You know, I can’t say for sure. I don’t remember specifically being offered [the option to speak with a genetic counselor before testing]. But it’s possible that I was offered it and overlooked it because I didn’t really feel that, you know, I had information gaps about genetic testing or genetic counseling. So I’m not sure if it was offered to me or not. But in either case, I would not have felt like I would have needed it.”*

An optional educational video e-mailed to participants prior to the result disclosure and genetic counseling session was not universally viewed. Of those who viewed the video, some reported that the information was helpful, while others suggested that the content may have been too complex.

> ***Participant 8 (age 65-69, unaffected female, negative results):*** *“I actually thought the video was very informative…I mean I don’t know what those genes are”*
>
> ***Participant 7 (age 65-69, unaffected male, negative results):*** *“I thought it was good. You know, it started out at a pretty simple level, but very quickly ramped up into some fairly sophisticated genetic information. I thought it was interesting. So I liked it.”*

A few participants suggested that having different forms of information presented (i.e. video and written material) was helpful, and one participant highlighted this fact by describing how they interacted with the informational video.

> ***Participant 2 (age 60-64, unaffected male, negative results):*** *“I was watching the video very carefully until I noticed that the wording of the video was somewhere else. Then I downloaded that. And I looked at the video. I was focused more on the written –- than the visual."*

In summary, receiving individual results was highly valued, with several participants expressing interest in obtaining additional personal research results; however, their preferences varied regarding the timing and way genetic information was communicated during the study process and counseling session.

### Theme 3: Emotional Experiences and Support Needs in Result Disclosure

Receiving genetic test results may evoke emotional responses in some individuals, and genetic counselors are trained to provide support during a counseling session [9]. When asked about their first reaction to their genetic results, many participants were shocked yet relieved results were negative, while those who tested positive seemed unsurprised.

***Participant 2 (age 60-64, unaffected male, negative results):*** *“[My reaction to the results] was surprise and then relief. Although, you know, I thought, but you’re not off the hook.”*

> ***Participant 13 (age 70-74, unaffected male, GBA1 positive and LRRK2 positive)*** *"I figured that they would probably be telling me that I had some markers. I would have been more surprised if there was nothing there"*

One participant was even disappointed with receiving negative results.

> ***Participant 5 (age 75-79, affected male, negative results):*** *“There was a part of me that was kind of rooting for being positive So I was probably a little disappointed that [the test] didn’t [show the reason I got Parkinson’s] but I was really happy that you guys are so thorough with the counseling.’ “*

As participants experienced a range of reactions to their results, they also recognized various supportive qualities of their genetic counselor and generally felt that they trusted the genetic counselor. One participant mentioned tone and empathy were important qualities.

> ***Participant 7 (age 65-69, unaffected male, negative results):*** *“the tone was empathic, caring and warm and genuine, you know, which is what you look for in terms of tonal qualities. And the content of what she said was very understanding, caring, inquiring as to whether we’re, you know, understanding what we’re talking about.”*

Even though participants appreciated a warm, empathic tone, when asked if emotional support was important, many stated that it was not needed during the session since their results did not evoke a particularly emotional response.

> ***Participant 5 (age 75-79, affected male, negative results):*** *“There was no need for me to cry on somebody’s shoulder, that kind of thing”*

However, most participants received negative results and several identified that emotional support could have been more important if their results had been positive. Interestingly, one unaffected participant with a positive genetic test result also did not feel a personal need for emotional support since the chance for PD was not 100%.

> ***Participant 4 (age 70-74, unaffected female, GBA1 positive result):*** *“if she’d given me bad news that I was guaranteed to get [Parkinson’s disease], then emotional support would’ve been appreciated. But since I’m not guaranteed, no, it was fine.”*

As shown, while participants experienced a range of emotional reactions to their genetic results and emphasized the importance of a supportive and empathetic genetic counselor, many indicated that emotional support for them personally was unnecessary.

### Theme 4: Communication Quality and Alignment with Participant Needs

Effective communication that aligns with participant needs is essential in genetic counseling to ensure understanding, foster trust, and provide appropriate support during a session [9]. Several participants reported that the counselor’s communication was clear and professional and delivered in a supportive way.

> ***Participant 2 (age 60-64, unaffected male, negative results):*** *“…clear straightforward. Concerned, but in an appropriate way, not in a creepy or unctuous kind of way”*

Participants reported appreciation for being listened to and for having the opportunity to ask questions during the session.

> ***Participant 5 (age 75-79, affected male, negative results):*** *“if there was something I didn’t understand or I wanted more information, I asked and she responded. So I thought it was a great session.”*

The desired level of involvement of participants in the communication process was variable. Some thought the genetic counselor was the “expert” and thought their role was the “listener”, whereas others wanted to be more involved in the communication process.

> ***Participant 2 (age 60-64, unaffected male, negative results):*** *“I think it was -- well, as you can tell, I’m pretty engaged as a person. So, yeah, -- there was lots of opportunities for me to be engaged and I felt like I had my questions answered”……… “it wasn’t like, you know, she said something and I said, oh, thank you and went away. I mean, it was a conversation and that’s what it should be.”*

However, on occasion there were mismatches in communication where participants were unclear about how or why certain words or phrases were used during the session.

> ***Participant 3 (age 75-79, affected female, LRRK2 positive result):*** *“There were one or two things that I was confused about. “And unfortunately, I didn’t ask her. I should have.” When she was talking about the LRRK2 changed and not changed. I wasn’t really sure about what that meant.”*

It was also noted in this instance that the communication of the genetic counselor outpaced the ability of the participant to fully grasp information or formulate and raise questions.

Overall, participants highlighted the value of clear, professional, and supportive communication in genetic counseling, appreciating being listened to and the ability to ask questions; however, differing preferences for involvement and occasional misunderstandings underscore the importance of tailoring communication to individual participant needs.

## DISCUSSION

This study adds to the limited research on participants’ detailed experiences with post-test genetic counseling and return of PD-related genetic results [10]. We found that personal connections to disease played a key role as a motivator for participation in the research; receiving individual results with as much information as possible was important; emotional reactions to results could not be entirely predicted; participants largely did not highlight a need for in-depth emotional support; and clear, empathic communication was highly valued. These findings align with the existing literature on return of results [10–15], while providing additional insights for researchers designing genetic studies, particularly in the neurodegenerative space.

The idea of personal connection permeated the interviews, suggesting that relational aspects of participation were central to individuals’ experiences. Participants commented that they felt heard, highlighting the role of human connection within the genetic counseling and research process. Personal or family experiences with disease frequently motivated participation and shaped engagement. Together, these findings suggest that incorporating personalized counseling and prioritizing human connection may enhance research engagement. This underscores the importance of intentionally designing genetic counseling and research interactions that allow time for individualized dialogue and relationship-building, thereby reinforcing participants’ sense of being valued contributors rather than passive research subjects. These observations align with other studies showcasing the importance of relationship [13–16]. Bardach et al [12] observed that personal connections to Alzheimer’s disease and positive relationships with the research team were key motivators for sustained research participation. Weemering et al. [13], in a systematic review and meta-analysis of trial participation in neurodegenerative diseases across multiple studies, found that 70% of respondents cited the relationship with staff as a primary reason for participation across multiple studies.

Participants reported that the most important information was learning their genetic test results. This matches literature showing that research participants desire return of genetic results and often want as much information as possible [15, 17,18]. Some participants desired results beyond what professionals typically prioritize, including additional study findings or more individualized information. Although not novel, this reinforces that offering research results, when feasible, may enhance perceived personal utility and support study enrollment.

Preferences for pre-disclosure education were variable. Most felt that formal pre-test genetic counseling should be offered but not required, and that educational resources should be available in multiple formats, such as written materials and videos. This reinforces that there is no one-size-fits-all approach to providing information [19]. Several participants were unaware of the option for pre-test counseling or had not viewed the informational videos, highlighting areas where improved strategies could increase access to educational and counseling resources.

Many participants expected positive results, which may reflect coping strategies or overestimation of personal risk prior to testing. Emotional responses ranged from surprise and relief to confirmation, consistent with prior studies demonstrating varied positive and negative reactions following genetic test disclosure [3, 10]. As reported previously in PD genetics research, reactions were not always anticipated, including disappointment with negative results [6, 20]. Overall, emotional responses were individualized and context dependent.

Trust, empathic communication, and support were universally valued by participants. However, the depth of emotional engagement appeared to vary according to study results, perceived risks, and expectations about the genetic counseling session. Radecki et al. [10] found that many PD research participants were unaware that emotional support could be part of genetic counseling, despite recognizing its potential importance. Although our participants did not express a strong need for emotional support, greater awareness of its availability may benefit those who do and may normalize its inclusion within a genetic counseling session.

Participants emphasized the importance of clear communication delivered in a supportive manner, consistent with core principles of genetic counseling as a relational practice grounded in empathy, trust, and respect [8,21]. Participants’ experiences suggest that effective genetic counseling in research settings involves conveying accurate information within a supportive interpersonal relationship.

Despite generally positive experiences, some participants described breakdowns in communication related to pacing or terminology. In some cases, participants were unclear about how or why certain words or phrases were used. This finding is consistent with prior research demonstrating that language used in genetic counseling is often confusing or misunderstood [22]. We agree with the viewpoint of Van Pottelberghe et al [23] that the terms used in genetics matter, impacting understanding of key concepts and how counseling is experienced. Those returning genetic results should use consistent, clear language [23]; explain unfamiliar terms; check understanding [24]; and tailor communication to individual preferences, values, backgrounds, and literacy [24,25]. Future research in this area could focus on the development of specific communication guidance and training for those doing disclosures in research settings, beyond what we provide here and beyond what has been created for clinical settings [24,25].

All participants were of Ashkenazi Jewish ancestry, which may limit generalizability to populations with less exposure to genetic testing and counseling. We interviewed fewer individuals with PD and fewer participants with positive results than anticipated, limiting characterization of these subgroups. Although the sample size was small, qualitative research prioritizes depth, and recurring themes supported analytic sufficiency guided by information power. Another potential limitation is using genetic counselors that could introduce the possibility of professional bias; however, our domain-specific knowledge, as research genetic counselors, also represents a methodological strength, enabling informed, contextually grounded analysis of complex counseling processes in the research setting.

Based on these findings, we provide guidance for researchers designing return-of-results processes (Table 2).

**Table 2.**
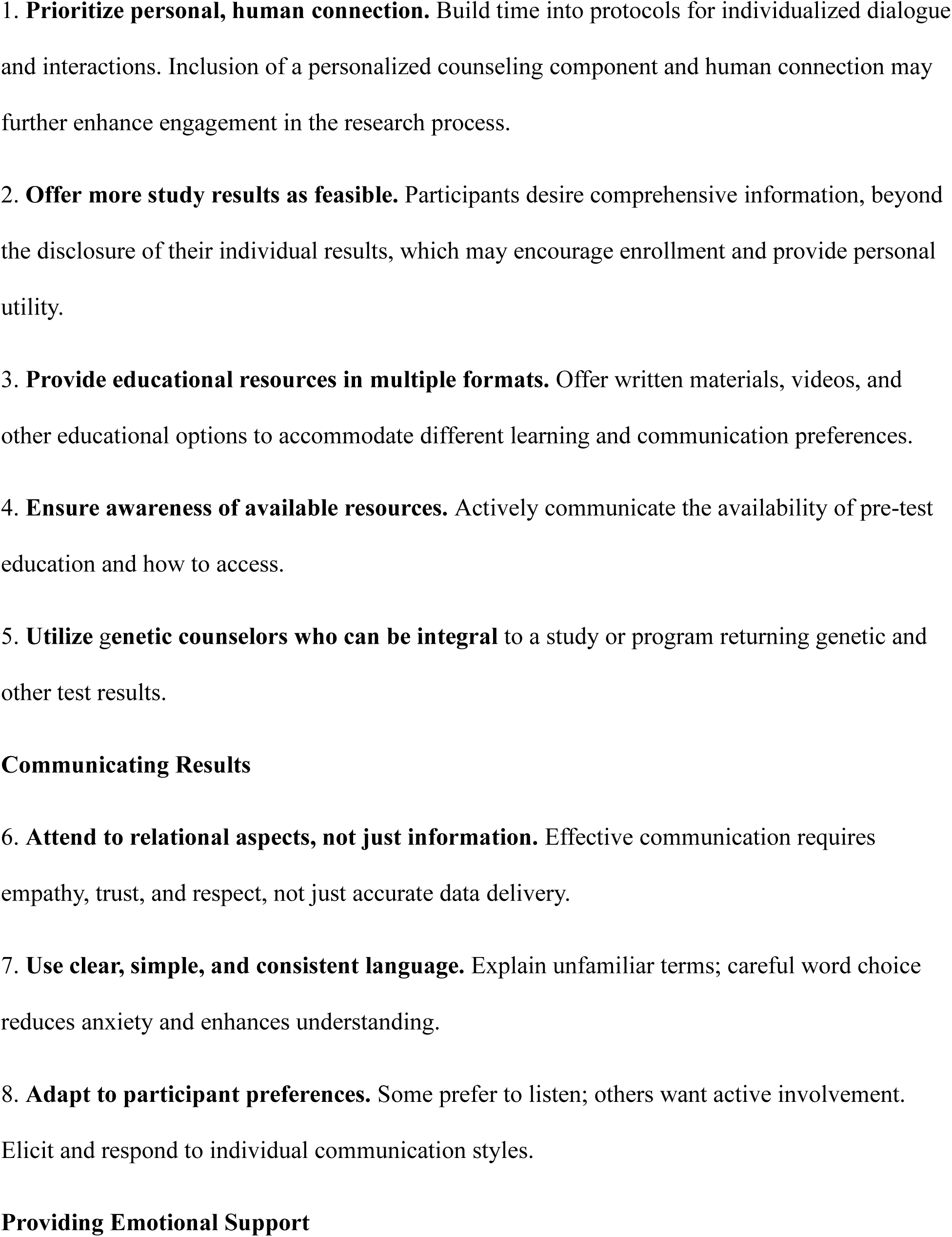
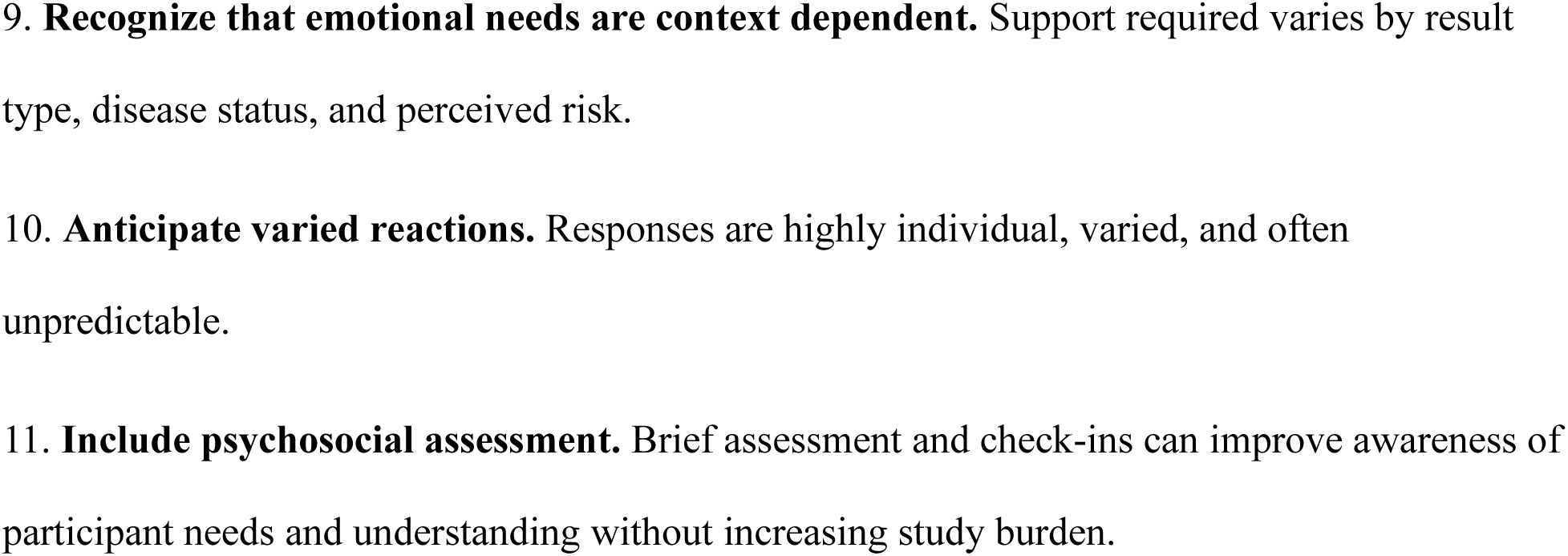
Returning Research Genetic Results: Key Considerations.

## Conclusion

In summary, this study provides additional detailed insights into the experiences of research participants receiving return of genetic results and post-test genetic counseling in the context of PD research. Our findings underscore that effective genetic test disclosures in research settings require attention to both the information provided and relational aspects. As genetic and biomarker testing become increasingly integrated into PD research and gene-targeted clinical trials expand, researchers and neurologists will need to thoughtfully design result disclosure processes that balance efficiency with personalization. This includes ensuring participants are aware of available educational resources, offering flexible formats for pre-test education with personalized information for post-test counseling, and recognizing that emotional needs vary considerably across individuals and result types. Future research should explore optimal strategies for tailoring genetic counseling based on participant characteristics, different populations, and types of results disclosed, as well as methods to enhance awareness and uptake of available support resources. Ultimately, integrating person-centered genetic counseling into research design may not only improve participant experiences but also strengthen research engagement and trust—essential elements as the field moves toward precision medicine for PD.

## Author Contributions

JV: Conceptualization, Data Curation, Formal Analysis, Methodology, Writing – original draft, Writing – review and editing; KF: Formal Analysis, Methodology, Writing – original draft, Writing – review and editing; LC: Conceptualization, Formal Analysis, Methodology, Writing – original draft, Writing – review and editing; MM: Conceptualization, Formal Analysis, Methodology, Writing – original draft, Writing – review and editing; KH: Conceptualization, Methodology, Supervising, Writing – review and editing. All authors read and approved the final manuscript.

## Acknowledgements

We thank all the individuals in PPMI who took the time and effort to be interviewed. Data used in the preparation of this article were obtained from the Widespread Recruitment Database (WRD). PPMI – a public-private partnership – is funded by the Michael J. Fox Foundation for Parkinson’s Research and funding partners, including Abbvie, Alamar Biosciences, Aligning Science Across Parkinson’s, Arrowhead Pharma, AskBio, BIAL, BioArctic, Biohaven, BlueRock Therapeutics, Bristol-MyersSquibb, Calico Labs, Capsida Biotherapeutics, Critical Path Institute, DaCapo Brainscience, Denali, Edmond J. Safra Foundation, Eli Lilly, Gain Therapeutics, GE HealthCare, Genentech, GSK, Insitro, Johnson & Johnson Innovative Medicine, Lundbeck, Merck, Neumora, Neuron23, Novartis, Regeneron, Roche, Sanofi, Tenvie, UCB, VanquaBio, Voyager Therapeutics, the Weston Family Foundation.

## Protocol Information

Protocol information for The Parkinson’s Precision Medicine Initiative (PPMI) Clinical – Establishing a Deeply Phenotyped PD Cohort can be found on protocols.io or by following this link: https://dx.doi.org/10.17504/protocols.io.n92ldmw6ol5b/v2.

## Declaration of use of generative AI and AI-assisted technologies in the manuscript

During the preparation of this work the authors used OpenEvidence and ChatGPT in order to assist with language editing and improving the clarity and readability of the manuscript. After using these tools, the authors reviewed and edited the content as needed and take full responsibility for the content of the published article.

## References

[1] Bloem, M.S. Okun, C. Klein, Parkinson’s disease, Lancet 397 (2021) 2284–2303. 10.1016/S0140-6736(21)00218-X .

[2] W. Eijzenga, N.K. Aaronson, D.E. Hahn, G.N. Sidharta, L.E. van der Kolk, M.E. Velthuizen, M.G. Ausems, E.M. Bleiker, Effect of routine assessment of specific psychosocial problems on personalized communication, counselors’ awareness, and distress levels in cancer genetic counseling practice: a randomized controlled trial, J. Clin. Oncol. 32 (2014) 2998–3004. 10.1200/JCO.2014.55.4576.

[3] C. Stracke, C. Lemmen, K. Rhiem, R. Schmutzler, S. Kautz-Freimuth, S. Stock, "You always have it in the back of your mind"—feelings, coping, and support needs of women with pathogenic variants in moderate-risk genes for hereditary breast cancer attending genetic counseling in Germany: a qualitative interview study, Int. J. Environ. Res. Public Health 19 (2022) 3525. 10.3390/ijerph19063525.

[4] T. Foroud, D. Smith, J. Jackson, J. Verbrugge, C. Halter, L. Wetherill, T. Sims, R. Xiao, C. Bhatti, Novel recruitment strategy to enrich for LRRK2 mutation carriers, Mol. Genet. Genomic Med. 3 (2015) 404–412. 10.1002/mgg3.151.

[5] K. Marek, S. Chowdhury, A. Siderowf, S. Lasch, C.S. Coffey, C. Caspell-Garcia, T. Simuni, D. Jennings, C.M. Tanner, J.Q. Trojanowski, L.M. Shaw, J. Seibyl, N. Schuff, A. Singleton, K. Kieburtz, A.W. Toga, B. Mollenhauer, D. Galasko, L.M. Chahine, D. Weintraub, The Parkinson’s progression markers initiative (PPMI)—establishing a PD biomarker cohort, Ann. Clin. Transl. Neurol. 5 (2018) 1460–1477. 10.1002/acn3.644.

[6] J. Verbrugge, L. Cook, M. Miller, M. Rumbaugh, J. Schulze, L. Heathers, L. Wetherill, T. Foroud, Outcomes of genetic test disclosure and genetic counseling in a large Parkinson’s disease research study, J. Genet. Couns. 30 (2021) 755–765. 10.1002/jgc4.1366.

[7] K. Malterud, V.D. Siersma, A.D. Guassora, Sample size in qualitative interview studies: guided by information power, Qual. Health Res. 26 (2016) 1753–1760. 10.1177/1049732315617444.

[8] V. Braun, V. Clarke, Using thematic analysis in psychology, Qual. Res. Psychol. 3 (2006) 77–101. 10.1191/1478088706qp063oa.

[9] B. Biesecker, Genetic counseling and the central tenets of practice, Cold Spring Harb. Perspect. Med. 10 (2020) a038968. 10.1101/cshperspect.a038968.

[10] M. Radecki, C. Halverson, L. Wetherill, M. Miller, Patient perceptions of genetic counselors’ role and emotional support needs in adults with Parkinson’s disease, J. Genet. Couns. 34 (2025) e1971. 10.1002/jgc4.1971.

[11] J.O. Robinson, J. Wynn, B. Biesecker, L.G. Biesecker, B. Bernhardt, K.B. Brothers, W.K. Chung, C.M. Christensen, J.R. Green, K.L. Lewis, S.A. Lyon, A.L. McGuire, Psychological outcomes related to exome and genome sequencing result disclosure: a meta-analysis of seven Clinical Sequencing Exploratory Research (CSER) Consortium studies, Genet. Med. 21 (2019) 2781–2790. 10.1038/s41436-019-0565-3.

[12] S.H. Bardach, K. Parsons, A. Gibson, G.A. Jicha, "From victimhood to warriors": super-researchers’ insights into Alzheimer’s disease clinical trial participation motivations, Gerontologist 60 (2020) 693–703. 10.1093/geront/gnz096.

[13] D.N. Weemering, A. Beelen, T. Kliest, L.H. Visser, L.H. van den Berg, R.P.A. van Eijk, Trial participation in neurodegenerative diseases: barriers and facilitators: a systematic review and meta-analysis, Neurology 103 (2024) e209503. 10.1212/WNL.0000000000209503.

[14] H. Webster, M. Zelinsky, V. Rico, K. Martinez, L. Sankary, Reconciling recommendations for ethical sharing of biomarker results with Alzheimer’s disease research participants: a systematic review and qualitative synthesis, J. Alzheimers Dis. (2025) 13872877251392555. 10.1177/13872877251392555.

[15] D.F. Vears, J.T. Minion, S.J. Roberts, J. Cummings, M. Machirori, M. Blell, M.J. Murtagh, Return of individual research results from genomic research: a systematic review of stakeholder perspectives, PLoS One 16 (2021) e0258646. 10.1371/journal.pone.0258646.

[16] J.M. O’Daniel, S. Ackerman, L.R. Desrosiers, S. Rego, S.J. Knight, L. Mollison, G. Byfield, K.P. Anderson, M.I. Danila, C.R. Horowitz, G. Joseph, G. Lamoure, N.M. Lindberg, C.K. McMullen, K.F. Mittendorf, M.A. Ramos, M. Robinson, C. Sillari, E.B. Madden, Integration of stakeholder engagement from development to dissemination in genomic medicine research: approaches and outcomes from the CSER Consortium, Genet. Med. 24 (2022) 1108–1119. 10.1016/j.gim.2022.01.008.

[17] J.M. Bollinger, J. Scott, R. Dvoskin, D. Kaufman, Public preferences regarding the return of individual genetic research results: findings from a qualitative focus group study, Genet. Med. 14 (2012) 451–457. 10.1038/gim.2011.66.

[18] D. Sheen, A. Willis, Z. Fehlberg, M. Bogwitz, S. Lunke, Z. Stark, C. Gaff, Systematic review of preferences for additional findings from genomic testing, Eur. J. Hum. Genet. 34 (2026) 10–26. 10.1038/s41431-025-01968-w.

[19] K. Fiallos, E. Selznick, J. Owczarzak, M. Camp, D. Euhus, M. Habibi, L. Jacobs, A. Johnson, C. Klein, J. Lange, P. Njoku, K. Visvanathan, Patient experiences of cancer genetic testing by non-genetics providers in the surgical setting, J. Genet. Couns. 34 (2025) e1983. 10.1002/jgc4.1983.

[20] M. Miller, L. Cook, J. Verbrugge, P.D. Hodges, K.J. Head, M.A. Nance, Delivering genetic test results for Parkinson disease: a qualitative approach to provider experiences in the PD GENEration study, Neurol. Clin. Pract. 14 (2024) e200282. 10.1212/CPJ.0000000000200282.

[21] J. Austin, A. Semaka, G. Hadjipavlou, Conceptualizing genetic counseling as psychotherapy in the era of genomic medicine, J. Genet. Couns. 23 (2014) 903–909. 10.1007/s10897-014-9728-1.

[22] A. Chapple, P. Campion, C. May, Clinical terminology: anxiety and confusion amongst families undergoing genetic counseling, Patient Educ. Couns. 32 (1997) 81–91. 10.1016/s0738-3991(97)00065-7.

[23] S. Van Pottelberghe, F.J. Hes, M. Genuardi, Advancing towards clear and patient-centred language in cancer genetics, Eur. J. Hum. Genet. (2025) Advance online publication. 10.1038/s41431-025-01976-w.

[24] G. Joseph, R. Lee, R.J. Pasick, C. Guerra, D. Schillinger, S. Rubin, Effective communication in the era of precision medicine: a pilot intervention with low health literacy patients to improve genetic counseling communication, Eur. J. Med. Genet. 62 (2019) 357–367. 10.1016/j.ejmg.2018.12.004.

[25] D.L. Cragun, M. Volesky, T. Scott, D. Cline, N. Khan, M. Holum, E. Fisher, A. Richardson, L. Clark, K.A. Binkowski, A. Imtiaz, H.A. Zierhut, Unveiling genetic counseling skills: developing an online training course and analyzing counselor communication practices, J. Genet. Couns. 34 (2025) e2002. 10.1002/jgc4.2002

