## Supplement for "More Than Results: A Qualitative Study on the Role of Person-Centered Genetic Counseling in Parkinson’s Disease Research"

QUALITATIVE STUDY TO EVALUATE GENETIC COUNSELING FOR PARKINSON'S DISEASE

INTERVIEW QUESTIONS

INTRODUCTION SCRIPT OUTLINE:

Hello my name is <name>, and I am a member of the Parkinson's Progression Markers Initiative (PPMI) research team at Indiana University and will be conducting this interview. I just wanted to remind you about a few details before we begin. The interview should take about 30-45 minutes to complete. I will be asking you questions about your genetic testing and counseling experience as a participant in the Parkinson's Progression Markers Initiative (PPMI) study. You are welcome to not answer any question and can stop the interview at any time. Please remember we are interested in your honest answers; we really want to know about your experiences so that we can improve our counseling sessions in the future. The interview will be recorded, are you ok with that? Are you ready to begin?

<u>Transition script:</u> These first set of questions will focus on learning about any previous experience you may have had with genetic counseling and testing.

1. Before our study, what had you **heard about genetic counseling**?

2. Had you ever **met with a genetic counselor** before?
    a. IF YES: Are you willing to share the reason?
        i. Tell me a little about that meeting.
            1. Was it a good or bad experience? Why?

3. Before this study, did you have any **other genetic testing**? (This could be with a clinic, like through your doctor, or with something like a direct to consumer service like 23andMe.)
    a. IF YES: What was that experience like?
        i. Would you say this a positive, negative, or neutral experience?
        ii. Why?

<u>Transition script:</u> Ok, let's move into some questions that are specific to your experiences with our Parkinson's study.

4. Let's start with talking about the **consenting process**. This was done at the beginning of the study. This process involved things like reviewing the information about the study on the website, reading the informed consent document, speaking with a study coordinator and possibly speaking with a genetic counselor before having testing.
    a. What did you think about the consenting process?
    b. Did you find it easy or difficult?
    c. What would you change about the process to make it better?

<u>Transition script:</u> Ok, let's focus on some other steps before genetic testing.

5. It is common practice for a genetic counselor to meet with a patient before having genetic testing. During a session like this, the genetic counselor would answer questions, review and discuss the information about the genetics of the condition, and review the risks, benefits and limitations of genetic testing. We call this **a pre-test genetic counseling session**. A pre-test genetic counseling session was not offered in this study, though participants could request to speak with a genetic counselor to have questions answered before having testing.
    a. If no pretest consult: Our records indicate you <u>did not</u> speak with a genetic counselor before having testing in this study.
        i. Were you aware this option was available?
        ii. Do you think a pre-test genetic counseling session would have been helpful for you?
            1. Why or why not?
            2. If you could go back, would you have chosen to do a pre-test session?
                a. What would you have asked during that session?
        iii. Do you think speaking to a genetic counselor before testing should remain optional or should a pre-test genetic counseling session be a standard part of the process?
            1. Why or why not?
    b. If pretest consult: Our records indicate you <u>did</u> speak with a genetic counselor before having testing in this study.
        i. Did you find speaking with a genetic counselor prior to having testing helpful or not helpful?
            1. Why or why not?
        ii. Do you think speaking with a genetic counselor prior to having testing should remain optional or should a pre-test genetic counseling session be a standard part of the process?
            1. Why or why not?

<u>Transition script:</u> Now we will move to some questions about a video we created for this study.

6. If you recall, we sent you an email confirming your genetic counseling appointment and we included a link to a video in the email. This video provided information about what to expect in the genetic counseling session and it outlined the genetics of Parkinson's disease. Some of the participants chose to watch the video before the genetic counseling session and some did not. Do you remember watching **the video**?
    a. IF NO: Why didn't you watch the video?
        i. Did you have any internet issues that prevented you from watching the video?
        ii. Would you have preferred the information in a different format, like a website with text or a pamphlet in the mail?
    b. IF YES: What did you think about the video?
        i. What did you like most or least about it?

ii. How many times did you watch the video?
iii. Did you watch the video before or after the genetic counseling session?
iv. Do you think watching the video earlier on in the study process, like during the consenting process, would have been helpful?
    1. Why or why not?

Transition script: Ok, now I'd like you to ask you some questions about the genetic counseling session when we shared your genetic testing results with you.

7. Before this call took place, what were you **expecting the genetic counselor to do or to talk about** during the session?

8. What was your **first reaction** when the genetic counselor shared your **genetic testing results** with you?
    a. Was there anything that surprised you about your test results?
        i. Do you remember if your results were negative or positive?
    b. How did your results make you feel at that time?
    c. Have your feelings about your results changed? If so, how?

9. What did you think about the education and information (**factual information**) that the genetic counselor provided during the genetic counseling session? (For example; she might have explained how a gene works, what a variant or a mutation is, how genetic factors are inherited and how they contribute to Parkinson's disease and the risk for relatives to develop Parkinson's disease.)
    a. What information in particular was helpful or not helpful?
    b. How or why was the information helpful or not helpful?
    c. Would you have wanted more or less education and information about these things during the call?
    d. Was there any part of the session with the genetic counselor that was different than what you had expected?

10. So, thinking about the general education and information that the genetic counselor provided you about genes and mutations, and the specific genetic testing results you received, did you feel like you **understood the meaning of your results at the close** of the genetic counseling session? If not, why?

11. Did you **share your results with anyone** after the call?
    a. Who? Why?
    b. How did those conversations go?
    c. What information did you share with them?

12. Now I want to ask a little bit about your impressions of the use of the telephone for providing genetic counseling. What did you think about using the **telephone mode** for receiving your results and having genetic counseling?

a. What did you like or not like about it?
b. If you had your choice, would you prefer to receive your results face to face, using a web-cam, or through telephone? Why would you prefer this method?

13. Was the genetic counseling **session too long or too short**, or just about right?
    a. Tell me more about that.

14. After the session, did you receive the **written information** from the genetic counselor in the mail or by email?
    a. If YES, what did you think about it?
        i. Was there any part of the materials that you think should be changed? How?

Transition script: Now we are going to focus on some questions about your genetic counselor. Remember we are interested in your honest experiences here, so please tell us how you really feel.

15. How would you describe the **genetic counselor's communication style**?
    a. For example, did they explain your results clearly?
    b. Were there any times where you had trouble understanding the information they shared with you?

16. Would you like to have been more or less involved in the session (ie doing the talking, guiding the info covered)

17. We know that receiving genetic testing results can be a scary and an emotional experience for some people, while it can also be empowering and bring clarity or answer questions for others. In these next couple of questions, we'd like to see how well you thought your genetic counselor did in **supporting you emotionally**.
    i. What did you like or not like about your genetic counselor?
    ii. Did you trust them?
        1. Why/why not?
    iii. Did you feel comfortable asking questions?
        1. Why/why not?
    iv. Did they seem to care about you?
        1. If yes, what made you think they cared.
        2. If no, what made you thinks they did not care.
    v. Did they offer any emotional support? If so, how? Was it enough?
    vi. Was receiving emotional support important to you? Why or why not?

18. Alright, thank you so much for sharing those impressions. We have a few **wrap up questions.**
    a. Overall, looking back at that phone call, what did you find most or least helpful about the genetic counseling telephone session?

19. Do you have any **recommendations** on how genetic counseling in this **study could be improved** for people with Parkinson's disease?
    a. If you could change anything about the genetic counseling you received, what would you change? Why?

20. Before we wrap up the interview, is there **anything else** you'd like to add about your genetic counseling experience?

Thank you for your time. We appreciate and value your feedback!
